# Understanding Covid-19 misinformation and vaccine hesitancy in context: Findings from a qualitative study involving citizens in Bradford, UK

**DOI:** 10.1101/2020.12.22.20248259

**Authors:** Bridget Lockyer, Shahid Islam, Aamnah Rahman, Josie Dickerson, Kate Pickett, Trevor Sheldon, John Wright, Rosemary McEachan, Laura Sheard, on behalf of the Bradford Institute for Health Research Covid-19 Scientific Advisory Group

## Abstract

**Background:** Covid-19 vaccines can offer a route out of the pandemic, yet initial research suggests that many are unwilling to be vaccinated. A rise in the spread of misinformation is thought to have played a significant role in this vaccine hesitancy. In order to maximise vaccine uptake it is important to understand why misinformation has been able to take hold at this time and why it may pose a more significant problem within certain populations and places.

**Objective:** To understand people’s Covid-19 beliefs, their interactions with health (mis)information during Covid-19 and attitudes towards a Covid-19 vaccine.

**Design and participants:** In-depth phone interviews were carried out with 20 people from different ethnic groups and areas of Bradford during Autumn 2020. Reflexive thematic analysis was conducted.

**Results:** Participants spoke about a wide range of emotive misinformation they had encountered regarding Covid-19, resulting in confusion, distress and mistrust. Vaccine hesitancy could be attributed to three prominent factors: safety concerns, negative stories and personal knowledge. The more confused, distressed and mistrusting participants felt about their social worlds during the pandemic, the less positive they were about a vaccine.

**Conclusions:** Covid-19 vaccine hesitancy needs to be understood in the context of the relationship between the spread of misinformation and associated emotional reactions. Vaccine programmes should provide a focused, localised and empathetic response to counter misinformation.

**Patient or public contribution:** A rapid community and stakeholder engagement process was undertaken to identify Covid-19 related priority topics important to both Bradford citizens and local decision makers.

## INTRODUCTION

Tackling the rise of misinformation, which we interpret as false or inaccurate information communicated irrespective of intent to deceive, is a central challenge of our age.^1^ The abundance of misinformation, facilitated by social media, has the potential for severe adverse consequences, as people become not only misinformed but less able to believe in scientific facts and trust experts.^2 3^ The rise and spread of misinformation is associated with periods of political and economic upheaval, and in the Covid-19 pandemic it poses a major threat to public health.^4^ Trust in government, scientists and health professionals is seen as essential in preventing the spread of Covid-19 and implementing a successful vaccine programme.^5 6 7^ Yet the spread of Covid-19 misinformation has contributed to what has been labelled as a ‘crisis of trust’.^8^ This decline in trust has been reinforced by legitimate criticism of government responses to the pandemic and the exacerbation of pre-existing mistrust in governments and health services, particularly amongst marginalised groups.^9 10 11^

There is evidence that fear and anxiety of catching and dying of Covid-19 is extensive, particularly by women, younger people and individuals who identified as being in recognised risk groups.^12^ As well as health anxiety, research has indicated that people are experiencing greater financial anxiety, loneliness and mental health issues as a result of government measures to prevent the spread of Covid-19.^13 14 15^ The constant news cycle around Covid-19 and the spread of misinformation is also reported to have exacerbated fear, anxiety and stress.^16 17^ Within this climate, the UK government is attempting to roll out a mass vaccination programme.

On the 2nd December 2020, the Pfizer/BioNTech Covid-19 vaccine was approved for use in the UK, with the first vaccine administered six days later. Initially focused on people over 80, people who live or work in care homes and health care workers at high risk, the vaccination programme is expected to be extended much more widely in 2021.^18^ The results from multiple surveys in the UK have found that between 54% and 64% of respondents would definitely or are very likely to accept a Covid-19 vaccine. Between 4% and 9% reported that they would definitely not or were unlikely to accept it, suggesting many people are unsure.^19 20 21^ There are already indications that certain population groups are more hesitant to receive a vaccine. A recent poll by the Royal Society of Public Health found that respondents from Black, Asian and minority ethnic (BAME) backgrounds were less likely to accept a Covid-19 vaccine (57%) compared to white respondents (79%).^22^

Vaccine hesitancy refers to delay in acceptance or refusal of vaccination despite its availability. This is not a new phenomenon, and has existed since vaccines were first introduced, and the reasons behind it are multifaceted and complex, with geographical variations. ^23 6^ In the last two decades, growth of vaccine hesitancy, particularly amongst new parents, has been assisted by a discredited study linking the MMR vaccine to autism and by the spread of misinformation and personal stories of alleged vaccine injury through the internet and social media.^24^ In the context of Covid-19, there is emerging evidence that people’s belief in misinformation about the virus and especially their views about the origin of Covid-19 (i.e. that it was manufactured) will make them less likely to accept a vaccine when it becomes widely available.^25 26^

Large scale surveys are helpful in identifying the general population’s intentions toward a Covid-19 vaccine and some barriers to uptake, including exposure to misinformation.^4 27^ Detailed qualitative work, however, enables us to further explore the interaction between misinformation and people’s experiences of and reactions to the pandemic, building understanding as to why vaccine hesitancy varies across populations and places. The aim of our study was to explore people’s Covid-19 beliefs, their interactions with health (mis)information during Covid-19 and attitudes towards a Covid-19 vaccine.

## METHODS

### Study design

This descriptive, inductive qualitative study was completed as part of a larger mixed-method, longitudinal research study to provide actionable intelligence to local decision makers, developed in response to community and stakeholder consultation processes described in Box 1. ^28^ We used in-depth interviews to explore citizen’s health experiences and beliefs during Covid-19. University of York ethical approval was secured in July 2020 (Ref: HSRGC/2020/400/G).

### Study setting

Our study was conducted in Bradford, a city in the North of England. Bradford and its surrounding district is the fifth largest metropolitan district in England and is an area of high deprivation and ethnic diversity. Since March 2020, Bradford has experienced a relatively high number of Covid-19 cases compared to the rest of the UK, and stricter lockdown measures from July 2020 which remained in place until the introduction of the tier system in October 2020.^29^ In the second wave of the pandemic, Bradford hospitals have experienced patient numbers similar to the peak of the first wave in April and May. High rates of Covid-19 in areas like Bradford are likely to be due to greater deprivation, high population density and a higher than average number of multi-generational households.^30^

### Sampling and data collection

We conducted in-depth interviews with 20 people in different communities and different areas of Bradford via a maximum variation sample, with our key sampling focus being diversity of ethnicity and age. Nine community influencers were contacted (three people from each major ethnic group in Bradford - South Asian, White British and Eastern European) and invited them to take part in an interview or identify others who would. This method was favoured because community influencers are trusted by their peers and people with whom they engage. Snowball sampling was used to recruit further participants. When 15 interviews had been completed, demographic and geographical gaps were identified, and additional participants were recruited via contact with volunteers at a community organisation. Conventional recruitment methods such as adverts on social media may not have attracted the diversity of respondents we were seeking.

Fieldwork took place between September and October 2020. Eleven women and nine men participated, ranging from 20 to 85 years old, but most were aged between 25 and 54 years. In terms of ethnic group, they identified as Asian or Asian British (Pakistani, Indian and Bangladeshi) (10) White British (6), White Other (Eastern European, Gypsy or Irish Traveller) (4). The participants lived in nine different Bradford postcodes, representing geographical and deprivation status spread. Half the participants were in paid or volunteer community roles and most tended to work in lower-paid occupations such as retail worker, carer or beauty therapist.

Due to social distancing measures, all interviews were conducted in English over the phone by the first author. Interviews in Urdu/Punjabi was available, but was not requested. The interviews ranged in duration from 30 to 90 minutes, with the average length being 55 minutes. All participants gave written, informed consent through one of the following methods: 1) taking a photo of their signed consent form and emailing it; 2) emailing or texting stating that they had read the information sheet and consent form and fully consented to taking part in the study or 3) sending a signed consent form via post or email. In addition, all participants confirmed consent verbally at the start of each interview. All interviews were digitally recorded and transcribed by a professional transcriber with identifying information removed and participants’ names pseudonymised.

### Interview questioning

Headline topic guide questioning was derived from the areas that the consultation process (Box 1) identified as important to explore. The format of the topic guide and interview questioning were flexible to allow participants to voice what they considered to be important. The initial topic guide contained open ended questions and was piloted and iterated several times particularly in relation to data generated in early interviews regarding people’s beliefs about a Covid-19 vaccine. We included all data in the analysis and did not discard meaningful data gathered during piloting.

### Analysis

We undertook the analysis using the principles of reflexive thematic analysis.^31^ Five interview transcripts, chosen based on their representativeness of the whole dataset were analysed independently by the first and last authors. We held an analysis session to identify commonalities and differences in the interview narratives and worked towards ordering the data into loose themes. The resultant coding framework was then refined further by the first author and applied to the rest of the dataset. The first author then coded all interviews and conducted further interpretive work to write up the findings, sense checking with the last author as necessary. The analysis conducted was manual without the use of a software package. The analysis was wholly inductive, and, as such, we did not structure it on any existing theoretical frameworks.

## FINDINGS

We will briefly set the context by describing participants’ personal experiences of Covid-19. None of our participants had ever had a positive Covid-19 test, although some suspected they might have had the virus in February/March before there was widely accessible testing. About a quarter of participants did not know anyone who had definitely had Covid-19. The remaining three quarters had friends, family members, colleagues or neighbours who had tested positive for Covid-19 and had experienced various symptoms and outcomes from being mildly ill to hospitalisation and even death. Finding out that someone they were close to had been very ill with Covid-19 did appear to make the virus more ‘real’.

The findings are presented in two sections: 1) *confusion, distress and mistrust*, which is a broad meta theme exploring participants’ narratives about their health beliefs during the pandemic and 2) *vaccine hesitancy and beliefs*, which was both a topic guide section but many participants spontaneously spoke about this during the interview. It is worth noting that the first broad theme is heavily linked to participants’ beliefs about whether they would accept or decline a forthcoming vaccine. That is, generally, the more confused, distressed and mistrusting participants felt about their social worlds during the pandemic – the less likely they were to be in favour of a vaccine for themselves or their families. We have not tried to separate out findings related to the concepts of confusion, distress and mistrust into three separate categories as it would be a false demarcation given they were so deeply interconnected.

### Confusion, distress and mistrust

#### Information and misinformation

The avalanche of information surrounding Covid-19 had left many of the people we interviewed feeling overwhelmed and confused. Participants reported accessing a wide variety of sources of information about Covid-19: television and radio (UK news stations and also news stations in Pakistan, India, Slovakia and Poland), online national newspapers (in the UK and elsewhere), local newspapers, the NHS website, the council website, YouTube, Facebook, WhatsApp, Twitter, Google and medical journals. Some participants said that they had made a decision to stop or limit their reading or watching of news about Covid-19 because it was too distressing and they could not make sense of it.

> *At the moment, I just, like distance myself from this, I don’t want to hear anything about it. (Sofija)*

When faced with seemingly contradictory information, they felt unsure about which sources could be trusted and what to believe, as this participant discusses:

> *There’s like all WhatsApp groups and things, there were just stuff flying around on that and videos and all sorts and it was just like awful, what is the truth, what’s not, how do you and it’s like I’ve always been a believer that obviously don’t watch YouTube, you know, don’t believe anything as such, you know, NHS, you know, your GP, your NHS, Government. Oh, it were all just contradicting. (Jackie)*

Participants felt the government response had been particularly bewildering. No-one we spoke to seemed to be staunchly anti-government and they expressed a desire to follow the rules and restrictions, but they also felt that the national government communication had been poor and their decisions were contradictory or hypocritical:

> *The government aren’t being clear and they’re saying one thing but then they’re saying other things, and basically what they’re trying to do, they’re trying to please everybody all of the time, it doesn’t happen. (Hasan)*

There was also mistrust for some traditional news outlets that people felt were a mouthpiece of the government. This state of confusion and mistrust was intensified for some people who were engaging with news sources from other countries. The governments in countries where some participants were born or that they had close connections to, such as Poland, Slovakia, Pakistan and India, were responding differently to the pandemic, and this affected how people viewed the UK situation:

> *Compared to Slovakia for example when they started, everybody were advised to wear mask, compared to the UK they weren’t encouraged to wearing masks, so they were contradicting each other, or who is telling us the best of truth. (Kristof)*

Some participants felt that more official health information should have been released in languages other than English, so that people could have understood the key messages better.

Amongst an array of conflicting information sources, social media stories about Covid-19 gained a lot of traction in Bradford (see Box 2). Whilst some of them were national or international, others had a more local focus. A number of participants said that they laughed at some of the stories that they encountered on WhatsApp and Facebook, however, the sheer volume of messages coupled with the fact that people they trusted were sharing them proved difficult to ignore. The conditions of the pandemic and lockdown, with people anxious and stuck at home, accelerated the spread of stories and the impact. As one person highlighted:

> *It got a bit too much on the internet, it was just too much, everywhere you look was just going, things going on, some are true, some are not true, so I kind of actually gave up looking on the internet, it put me off it because it was literally too overwhelming and you’re already stuck inside… you don’t know if it’s true and then it just makes you more scared half the time. (Ambreen)*

Participants underlined how quickly social media stories were shared, with Tariq stating “they just forward it straight away and then it just spreads like wildfire”. The more controversial or dramatic the posts or videos were, the more they spread. Individuals in these videos were (or were posing as) trusted professionals, such as a teacher, nurse or doctor. Being able to deliver a video in multiple languages indicated higher levels of education and trustworthiness.

> *They’re just so passionate the way they talk, they grab your attention and they’ve got you and the way they’re speaking and the terminology they’re using and they give you the facts and the figures and then you just get drawn and locked into it. (Tariq)*

When participants talked about their interactions with false news they distanced themselves, referring to them as something they were dismayed at or amused by, scrolled past or ignored. Yet their narratives revealed much more complex responses. Participants described the dilemma of not knowing what to trust or who to listen to, meaning they could not dismiss these stories entirely. They knew conspiracy theories and fake news existed but found it difficult to separate it from legitimate information, especially if it was being constantly repeated. Masood, for example, was very keen to stress he had a scientific education and read medical journals, but he spoke of feeling confused after reading something online which made him wonder whether the virus was man-made.

Stories shared were frequently very emotive, catching participants at a particularly anxious time when they were more willing to believe them. Louise described spending the whole day crying after watching a video which stated that on a particular day, ambulances would not be dispatched for asthmatic patients and people would be left at home to die. Louise’s son had asthma and before she found out it was ‘fake’ she had already shared the video amongst her family and friends because she was so upset and concerned. This gives some insight into how and why videos like this spread. Laila referred to a story about children being forcibly taken out of school and quarantined away from their parents:

> *So many people were talking about it and the way that the video was made it was like a proper…it convinced you. So I think everybody believed it. But then afterwards they said it was fake news. But by the time you find out the video was fake, you already believed it, you’ve stressed yourself out already. (Laila)*

Rapid local and targeted responses appeared to stem the tide of misinformation to some degree. Participants who discussed the school children story also talked about the video from the council which debunked the story and was produced in Urdu and Punjabi to reach the population where it had spread the most. In general, people trusted their local leaders and the council website was frequently mentioned as a place where people accessed information about Covid-19.

#### Vaccine beliefs and hesitancy

A local Bradford survey indicated a higher level of vaccine hesitancy than the UK as a whole. Out of 222 people, only 34% said they would definitely have it and 11% did not want it.^32^ Of the 20 people we interviewed for this study, nine were happy to have a Covid-19 vaccine (with caveats around safety), five felt very mixed, and six said that they would not be willing to have it. Generally, the people we spoke to were relatively positive about vaccinations in general with most having been immunised as children and having their own children immunised. The findings below offer some insight into how exposure to misinformation during Covid-19 may have made people particularly unwilling or hesitant to have the vaccine.

#### Safety concerns

The safety of a potential Covid-19 vaccine was a concern, even for those who were very willing to have it. Some felt reassured by the medical establishment testing process in the UK. Angela commented “in England we’re very good at testing stuff, aren’t we?” Louise was less sure:

> *I think I’d have to know that it was a safe*… *I mean, they wouldn’t be doing an unsafe vaccine anyway would they, you know, but I think I’d have to have some confidence that it was a good vaccine and that it was quite safe. (Louise)*

A major issue for people was how quickly any potential vaccine would have been produced, and that the vaccine makers would not know all the side effects as yet. Sofija was worried it had not had time to be fully tested and Tariq wanted to wait three to six months to see what the effects of the vaccine were on others before he would be happy to take it. Some participants were afraid of very severe side effects, and it was clear that these worries had been exacerbated by engagement with social media stories:

> *People are saying they don’t know how safe it is, plus they’ve made it so quick we don’t know the side-effects it’s going to have in the future. I mean it’s probably safe because they wouldn’t be allowed obviously to give it to us otherwise, or maybe they would you know, sometimes they don’t care, but you just don’t know if it could cause infertility, it could cause cancer in the future. (Rebecca)*

#### Negative stories and misinformation focusing on the vaccine

Those hesitant about having the vaccine felt confused by the negative stories about it, rather than being resolutely against it. There was one exception, Faiza, who had joined live social media broadcasts where people were revealing the ‘truth’ about the negative side effects of the vaccines that they said was being hidden from the general public. Other people’s engagement with misinformation around the vaccine was more passive. Rebecca did not actively seek out these types of videos but said they often auto-played when she was watching YouTube and she described feeling confused after watching one. Tariq knew that watching these videos was impacting on his feelings about the vaccine and vouched to stop watching them. Alongside videos which claimed that the Covid-19 vaccines were unsafe, there were also rumours that certain communities and ethnic groups were being targeted to test the vaccine, or the vaccine was being used as a way to harm them:

> *I think what the community are saying is that the vaccine is testing people, they’re just using people as the guinea pigs… we experience discrimination for many years, and if we’ve been focused for, if the Slovakian authorities we are focused especially on the Roma, and the focus is they will be testing them because they were noting who could be spreading all this coronavirus, they may think the same thing now why are we going to offer immunisation, because they’re going to trial it out on us. (Kristof)*

Angela was keen to have the vaccine and get on with normal life but indicated that she gave some credence to a story she had heard about people being injected with the Covid-19 virus instead of the seasonal flu vaccine. These examples demonstrate the way that misinformation has the potential to affect participants’ viewpoints in a myriad of divergent ways.

#### Personal Knowledge

A lot of the hesitancy around Covid-19 vaccine was rooted in lay beliefs about health, disease and vaccines, which appeared to have been influenced by recent exposure to misinformation. A view widely held, even by those participants positive about the Covid-19 vaccine, was that the seasonal flu jab can give a patient the flu. For some participants, these fears about the seasonal flu vaccines were transferred to a potential Covid-19 vaccine as it was regarded as something which would disrupt the body’s natural state:

> *If your body reacts to it it’s okay, if it can take it, your body fights, rejects it then what’s going to happen then? You know, so some people their bodies asking whether they can take it, some people’s bodies are weak, the immune system might not be working all that perfectly, or all that well, or that that might cause them a harm, instead of doing good. (Atif)*

Several participants showed a partial understanding of disease transmission and vaccinations but often had contradictory narratives. This only served to exemplify the complexity of the topic, which was made more confusing through frequent subjection to misinformation. Another perception about the Covid-19 vaccine was that it would be ‘stronger’ and involve a higher dose than other vaccines, which would be dangerous for recipients.

## DISCUSSION

This study aimed to explore the health beliefs of citizens in Bradford regarding Covid-19, alongside their attitudes towards a vaccine. We found that participants encountered a range of misinformation, usually through social media sources. This led to confusion, distress and mistrust in participants’ everyday lives and beliefs about government institutions and health services, a more general phenomenon.^33^ Vaccine hesitancy could be attributed to: safety concerns, negative stories and personal knowledge, all of which had been amplified by recent exposure to misinformation via social media. We found that the more confused, distressed and mistrusting the participants felt during Covid-19, the more likely they were to be hesitant about uptake of the Covid-19 vaccine.

In February 2020, the World Health Organization stated that the Covid-19 outbreak and response has been accompanied by a massive ‘infodemic’.^34^ The abundance of information about Covid-19 transmitted across a multitude of platforms has continued, making it difficult to discern what is accurate and what is not.^35^ We do not yet know exactly how far-reaching and impactful misinformation about Covid-19 has been or will be, but early evidence suggests that susceptibility to misinformation about Covid-19 negatively affects people’s willingness to get vaccinated.^4^This presents a global public health challenge.^36^ Our findings offer insight into why misinformation has taken a particular hold during this time and in certain places.

We know from previous research that misinformation thrives in times of stress and uncertainty, and Covid-19 has provided a perfect breeding ground, on both a global and local scale. ^4 37 38^ Research has also indicated that feeling anxious makes people more willing to believe misinformation even if it is inconsistent with their world view.^39^ Our participants’ narratives conveyed this sense of anxiety and insecurity, as well as the additional stress of seeing conflicting information and holding contradictory viewpoints. This appeared to be particularly heightened within marginalised groups, who had pre-existing reasons for mistrusting institutions and felt more at risk from the virus.

## IMPLICATIONS FOR POLICY AND PRACTICE

Local decision makers have the ability to counter misinformation by implementing targeted local responses. To do this successfully and quickly, there has to be systematic monitoring of the circulation of misinformation on social media. Participants’ trust in national government, health professionals and NHS organisations appeared to have weakened over the duration of the pandemic but there was evidence that they did trust people within community support roles that they had frequent contact with e.g. teachers, nursery workers, and advice workers. Effectively harnessing these connections, through trusted community networks and providing information in languages spoken locally, will be central to ensuring the spread of correct information and providing reassurance.

Hesitancy around the Covid-19 vaccine appears to be rooted in anxiety fuelled by misinformation and there is a need for this to be mediated by clear, honest and responsive information that is sensitively framed and non-judgemental. It would be prudent for health, social and community workers to be provided with an updated summary of locally circulating misinformation with helpful resources to help them counter concerns and provide informed reassurance.

### LIMITATIONS

Interviews took place before announcements about efficacious Covid-19 vaccines were made in November 2020. Participants’ discussions about a Covid-19 vaccine were therefore hypothetical. We do not know how these announcements, and the subsequent surge of information and misinformation, will have impacted on acceptability of a Covid-19 vaccine.

The research was conducted in one place with specific population demographics, and may, therefore, not be widely generalisable. However, there are places all over the UK which have multi-ethnic communities, similar levels of deprivation and population density, and have experienced comparable rates of Covid-19 cases and deaths, which we predict will have similar problems with misinformation spread.

## CONCLUSION

Our study found there is an intensity of misinformation being spread about Covid-19 in Bradford and this has impacted on participant’s lives by evoking confusion, distress and mistrust during the pandemic. Heightened levels of confusion, distress and mistrust are related to a lower proclivity towards Covid-19 vaccine uptake. As is often found with the inverse law of care, the people most likely to be affected by Covid-19 are those who are most hesitant towards the vaccine. Of critical importance to decision makers is the ability to understand misinformation in its local context and countering it in a sensitive and non-judgemental way via trusted local people whose opinion is valued in their community.

## Data Availability

The data are not publicly available due to privacy or ethical restrictions. Requests, to the corresponding author, for access to the data underpinning this paper will be considered and accommodated where reasonable.

## CONFLICTS OF INTEREST

The authors declare no conflicts of interest.

## FUNDING INFORMATION

▪ The Health foundation Covid-19 Award (2301201)
▪ A Wellcome Trust infrastructure grant (WT101597MA)
▪ The National Institute for Health Research under its Applied Research Collaboration Yorkshire and Humber (NIHR200166)
▪ ActEarly UK Prevention Research Partnership Consortium (MR/S037527/1)

## ACKNOWLEDGEMENTS

The authors would like to thank Professor Neil Small, Dr Sufyan Dogra, Dr Jennifer Hall, Professor Claire Bambra and Dr Louis Goffe for their advice and guidance in the development of the interview questions. We are grateful for all the support and feedback given to us by members of the Bradford Covid-19 Scientific Advisory Group. Finally, we would like to thank all our interview participants who took the time to be part of our study in a difficult year.

## Box 1

**PPI and stakeholder consultation process to identify critical topics**

In March 2020, Bradford’s Covid-19 Scientific Advisory Group was formed to support policy and decision makers in Bradford and the UK to deliver an effective urgent response and to better understand the wider societal impacts of Covid-19. As part of these aims, a rapid community and stakeholder engagement process was used to identify priority topics important to both citizens in Bradford and local decision makers. This process took place in April 2020 via the following engagement activities:

1. BL spoke to nine members of Bradford’s District Gold Command (established in response to the Covid-19 emergency). These were brief 15-20 minute phone calls to assess their top knowledge Covid-19 priorities.
2. Analysis of the first 350 free text responses to the Born in Bradford Covid-19 adult questionnaire. This survey was undertaken during the initial weeks of first lockdown to assess what the main concerns were for parents in the district.
3. SI and AR collected soft intelligence from 13 people considered influential within diverse community settings.

From the above, three priority topic areas were identified: 1) “health beliefs” which is the subject of this paper and encompassed: access to healthcare services, experiences of Covid-19, sources of health information and the spread of misinformation 2) adolescent mental well-being and 3) people living in poverty before Covid-19. Soft intelligence work continued throughout the development of the study and indicated that misinformation around a potential Covid-19 vaccine was gaining a lot of traction within Bradford. As a result, interview questions were shaped to reflect this and include questions about vaccine attitudes.

## Box 2

**Social media Covid-19 stories discussed by participants National/International**

▪ Covid-19 is not real, it is an effort to control society
▪ Covid-19 has been manufactured by China or other governments for control purposes
▪ Covid-19 is caused by 5G
▪ Covid-19 has been invented to make people use contactless payments so that the government can track individuals
▪ The Covid-19 vaccine contains a chip that will track individuals, stop them travelling etc.
▪ The Covid-19 vaccine will make people infertile and is an attempt to reduce the population, particularly targeted at people from BAME communities
▪ Covid-19 testing gives so many false positives that it is ineffective and you should not self-isolate
▪ Covid-19 exists but is not as virulent as the government says it is

**Regional/Local**

▪ If children test positive for Covid-19 during school hours they can be taken away into care and will not be able to see their parents until they test negative
▪ Health professionals at Bradford Royal Infirmary were injecting people with the Covid-19 virus, or killing people with Covid-19
▪ Bradford Royal Infirmary were inflating the numbers of people with Covid-19
▪ The health service was so overwhelmed that ambulances would not arrive in an emergency

## Notes

### Competing Interest Statement

The authors have declared no competing interest.

### Clinical Protocols

https://wellcomeopenresearch.org/articles/5-191/v1

### Funding Statement

This study was supported by the following research funding: The Health foundation Covid-19 Award (2301201). A Wellcome Trust infrastructure grant (WT101597MA). The National Institute for Health Research under its Applied Research Collaboration Yorkshire and Humber (NIHR200166). ActEarly UK Prevention Research Partnership Consortium (MR/S037527/1)

### Author Declarations

University of York ethical approval was secured in July 2020 (Ref: HSRGC/2020/400/G).

